# Metabolic Subphenotypes of Obstructive Sleep Apnea: NHANES 2017-2020 (pre-pandemic)

**DOI:** 10.1101/2025.08.01.25332658

**Authors:** Kaixuan Fu, Jinxiu Zhang, Xuefeng Zhu, Jikai Song, Bowen Xu, Haibin Dong, Jun Yang, Chunxiao Wang, Lei Gong, Lin Zhong

## Abstract

**Background:** OSA and MetS have a bidirectional relationship but increasing evidence suggests metabolic heterogeneity in OSA, systematic phenotyping of metabolic drivers in OSA are lack.

**Objective:** To identify metabolic subphenotypes of OSA and elucidate potential pathophysiological mechanisms using population-level data.

**Methods:** To analyze the data related to OSA and MetS from 2,260 participants in the NHANES database (2017–2020, pre-pandemic) using PCA and machine learning.

**Results:** 48.74% (1689/3465) participants were identified as OSA. PCA revaled that the first 6 PCs explained 85% of the variance in both overall and OSA participants, encompassing indicators of obesity, hypertension, dyslipidemia, and IR. Indicators with |loading value|≥0.6 in each PCs included obesity (Height, Weight, BMI, Waist, Hipline, W/H), blood pressure, blood lipids (CHOL, TG, LDL, HDL), and IR (GHB, GLU, VAI, LAP, TyG). Cluster analysis divided the overall participant into 6 clusters. Compared with Cluster 1 (32.92%), Cluster 2 (57.04%), Cluster 3 (57.21%), Cluster 4 (61.94%), Cluster 5 (47.69%) and Cluster 6 (50.80%) represented groups with higher prevalence of OSA, characterized by IR, isolated obesity, central obesity combined with hypertension, dyslipidemia and hypertension respectively. OSA participants were divided into 8 clusters. Cluster A, Cluster C, Cluster E, Cluster F were characterized by hypertension, dyslipidemia, isolated obesity and obesity combined with hypertension respectively. Cluster B had metabolic indicators better than average level. Clusters G and H were mainly characterized by IR.

**Conclusion:** Metabolic heterogeneity in the population is associated with the incidence of OSA. Metabolic characteristics of OSA populations may guide the treatment of OSA and its comorbidities.

## Instruction

Obstructive sleep apnea (OSA), a common sleep disorder, is characterized by partial or complete obstructed upper airway during sleep, leading to recurrent episodes of apnea (breathing pauses) or hypopnea (reduced breathing)[1]. Epidemiological research has demonstrated that OSA is highly prevalent, affecting approximately 10% of the population domestically.[2]. Apart from daytime sleepiness, memory impairment and cognitive decline, the pathophysiological mechanisms of OSA can cause chronic intermittent hypoxia (CIH), oxidative stress, heightened sympathetic nervous system activity, and systemic inflammation[3]. These alterations contribute to various chronic diseases, including cardiovascular diseases, diabetes, and chronic obstructive pulmonary disease (COPD)[3].

Metabolic syndrome (MetS) is a pathological condition characterized by obesity, hypertension, insulin resistance (IR), and hyperlipidemia[4]. Growing evidence suggests a bidirectional relationship between MetS and OSA[5]. Epidemiological studies have shown that MetS is highly prevalent among patients with OSA, and the risk of developing MetS increases with the severity of OSA[5]. Conversely, the presence of MetS may exacerbate OSA symptoms, thereby creating a vicious cycle [6].

However, the relationship between OSA and MetS involves more complex interactions. Individual components of MetS may independently influence OSA through distinct mechanisms. Leveraging NHANES data, our study focuses on dissecting the independent contributions of each MetS criterion to OSA using machine learning methods. This approach aims to identify key metabolic drivers of OSA, offering insights for targeted interventions in this high-risk population.

## Method

### Study Population

The National Health and Nutrition Examination Survey (NHANES) is a national, stratified, multi-stage probability sampling survey conducted by the National Center for Health Statistics (NCHS) to assess the relationship between nutritional health and disease prevention. The survey combines interviews and physical examinations, covering multiple aspects such as demographics, physical examination, laboratory tests, and questionnaires. More information about the database could be visited on [NHANES website](http://www.cdc.gov/nhanes).

Between 2017 and 2020 (before the COVID-19 pandemic), a total of 15,561 participants were included in the survey. Based on strict inclusion and exclusion criteria, this study ultimately identified 2,260 U.S. adults from the 2017-2020 NHANES cycle as the sample. Specifically, we excluded 6,369 participants lacking data on OSA, 5,930 participants lacking data on physical examination, or laboratory tests, 493 participants aged under 18 or over 65 years, and 509 participants lacking covariate data (Figure 1).

**Figure 1.**
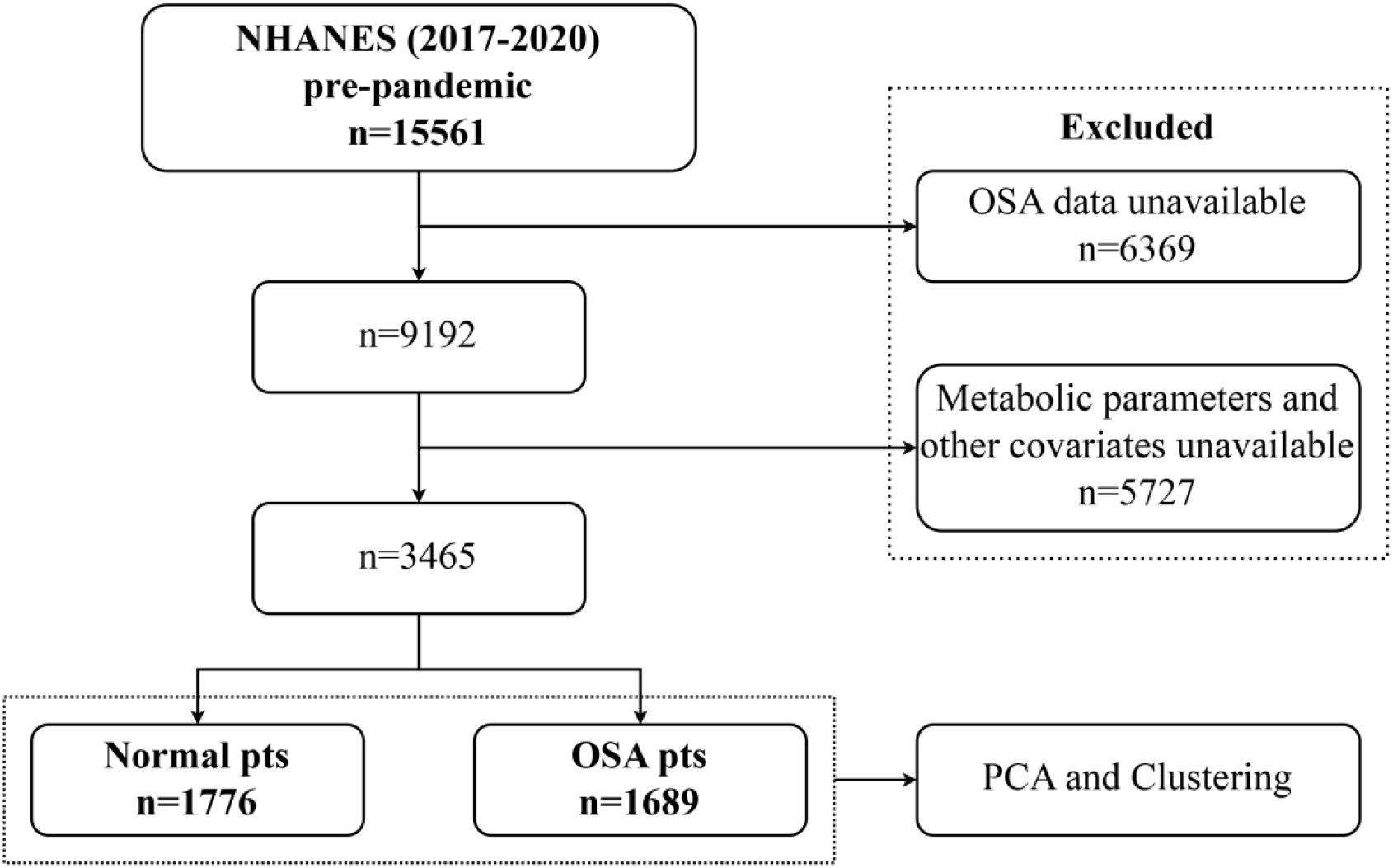
Flowchart of participants screening.

### Assessment of OSA

In accordance with other studies[7, 8] OSA was dignosed on the Questionnaire Data: (1) being excessively sleepy in the day even though they get at least 7h of sleep per night, as reported 16-30 times; (2) experiencing episodes of gasping, snorting, or stopping their breath on three or more occasions per week; (3) snoring on three or more occasions every week. When one or more answers of the qustions were “yes”, the participant was identified as OSA.

### Assessment of metabolic indicators and other covariates

#### Metabolic Indicators

Metabolic indicators include obesity indicators [Height, Weight, BMI, Waist, Hipline, waist/hipline ratio (W/H)], with length indicators measured in “meter (m)” and weight indicators measured in “kilogram (kg)”. Blood pressure indicators (SBP, DBP, MAP), all measured in “mmHg”. Hyperlipidemia indicators [cholesterol (CHOL), triglycerides (TG), low-density lipoprotein (LDL), high-density lipoprotein (HDL)], all measured in “mmol/L”. IR indicators [glycated hemoglobin (GHB, measured in “mmol/L”), fasting blood glucose (GLU, measured in “mmol/L”), LAP, TyG, VAI][8]. The TyG index was computed by using the following formula: Ln [fasting triglycerides (mg/dl)* fasting blood glucose (mg/dl)/2][9]. LAP and VAI were calculated according to gender using the following formulas in Supplementary Figure 1, where TG and HDL are in mmol/L, Waist is in cm, and BMI is in kg/m^2^[10].

#### Other Covariates

Age, race, gender, educational attainment, marital status, and the income-to-poverty ratio among the demographic factors were analysized with metabolic indicators.

### Statistical analyses

In accordance with NHANES guidelines, the analysis in this study employed interview weight variables (WTINT2YR) to ensure that the results were representative at the national level. To properly account for the standard errors arising from the complex sampling design, the analysis incorporated both the primary sampling unit variable (SDMVPSU) and the stratification variable (SDMVSTRA).

Continuous variables are summarized as the means with 95% confidence intervals (95% CIs) or medians with interquartile ranges, depending on the variable distribution, whereas categorical variables are depicted as counts and proportions. Based on the normal or non-normal distribution of the data and homogeneity of variance, t-tests, ANOVA, and non-parametric tests were employed to compare differences between groups. Z-score standardization was applied prior to principal component analysis (PCA) to eliminate scale differences. Principal components (PCs) that collectively explained 85% of the variance were retained for further analysis. For each retained PC, variables with |loading values| >0.6 were selected as the basis for cluster analysis. Radar charts were plotted using max-min normalized data for each variable within clusters. A p-value <0.05 was considered statistically significant. All analyses and visualizations were performed using SPSS 29.0, GraphPad Prism 10.0, OriginPro 2025, and Python 3.12.4.

## Results

### Baseline Characteristics of Participants

The baseline characteristics of the 3465 participants are presented in Table 1. Among them, 1689 participants (48.74%) were identified as OSA participants. Except for race and income-to-poverty ratio, all variables showed statistically significant differences between OSA and Normal participants.

**Table 1.**
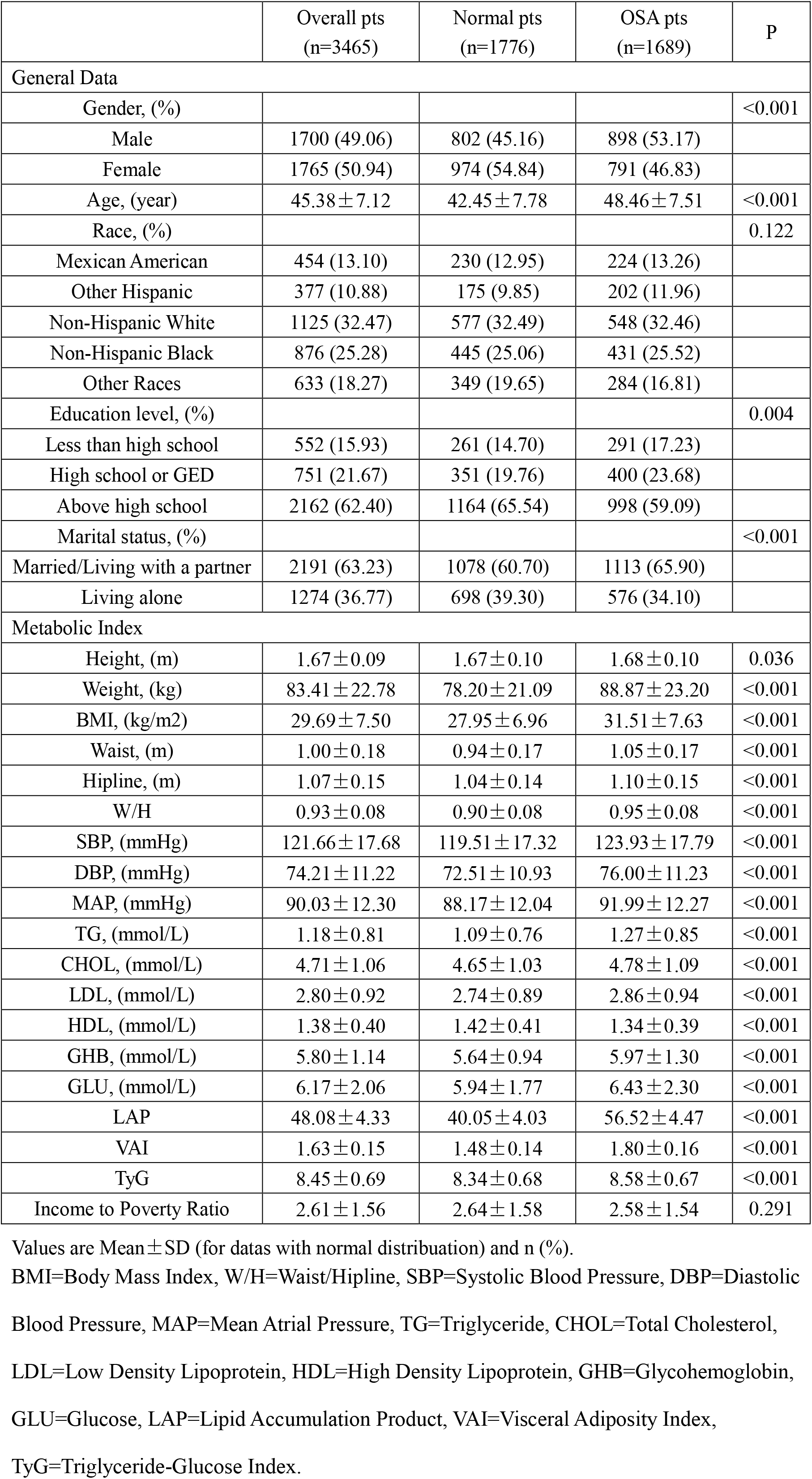
Baseline characteristics of all participants.

|  | Overall pts<br>(n=3465) | Normal pts<br>(n=1776) | OSA pts<br>(n=1689) | P |
| --- | --- | --- | --- | --- |
| General Data |  |  |  |  |
| Gender, (%) |  |  |  | <0.001 |
| Male | 1700 (49.06) | 802 (45.16) | 898 (53.17) |  |
| Female | 1765 (50.94) | 974 (54.84) | 791 (46.83) |  |
| Age, (year) | 45.38±7.12 | 42.45±7.78 | 48.46±7.51 | <0.001 |
| Race, (%) |  |  |  | 0.122 |
| Mexican American | 454 (13.10) | 230 (12.95) | 224 (13.26) |  |
| Other Hispanic | 377 (10.88) | 175 (9.85) | 202 (11.96) |  |
| Non-Hispanic White | 1125 (32.47) | 577 (32.49) | 548 (32.46) |  |
| Non-Hispanic Black | 876 (25.28) | 445 (25.06) | 431 (25.52) |  |
| Other Races | 633 (18.27) | 349 (19.65) | 284 (16.81) |  |
| Education level, (%) |  |  |  | 0.004 |
| Less than high school | 552 (15.93) | 261 (14.70) | 291 (17.23) |  |
| High school or GED | 751 (21.67) | 351 (19.76) | 400 (23.68) |  |
| Above high school | 2162 (62.40) | 1164 (65.54) | 998 (59.09) |  |
| Marital status, (%) |  |  |  | <0.001 |
| Married/Living with a partner | 2191 (63.23) | 1078 (60.70) | 1113 (65.90) |  |
| Living alone | 1274 (36.77) | 698 (39.30) | 576 (34.10) |  |
| Metabolic Index |  |  |  |  |
| Height, (m) | 1.67±0.09 | 1.67±0.10 | 1.68±0.10 | 0.036 |
| Weight, (kg) | 83.41±22.78 | 78.20±21.09 | 88.87±23.20 | <0.001 |
| BMI, (kg/m<sup>2</sup>) | 29.69±7.50 | 27.95±6.96 | 31.51±7.63 | <0.001 |
| Waist, (m) | 1.00±0.18 | 0.94±0.17 | 1.05±0.17 | <0.001 |
| Hipline, (m) | 1.07±0.15 | 1.04±0.14 | 1.10±0.15 | <0.001 |
| W/H | 0.93±0.08 | 0.90±0.08 | 0.95±0.08 | <0.001 |
| SBP, (mmHg) | 121.66±17.68 | 119.51±17.32 | 123.93±17.79 | <0.001 |
| DBP, (mmHg) | 74.21±11.22 | 72.51±10.93 | 76.00±11.23 | <0.001 |
| MAP, (mmHg) | 90.03±12.30 | 88.17±12.04 | 91.99±12.27 | <0.001 |
| TG, (mmol/L) | 1.18±0.81 | 1.09±0.76 | 1.27±0.85 | <0.001 |
| CHOL, (mmol/L) | 4.71±1.06 | 4.65±1.03 | 4.78±1.09 | <0.001 |
| LDL, (mmol/L) | 2.80±0.92 | 2.74±0.89 | 2.86±0.94 | <0.001 |
| HDL, (mmol/L) | 1.38±0.40 | 1.42±0.41 | 1.34±0.39 | <0.001 |
| GHB, (mmol/L) | 5.80±1.14 | 5.64±0.94 | 5.97±1.30 | <0.001 |
| GLU, (mmol/L) | 6.17±2.06 | 5.94±1.77 | 6.43±2.30 | <0.001 |
| LAP | 48.08±4.33 | 40.05±4.03 | 56.52±4.47 | <0.001 |
| VAI | 1.63±0.15 | 1.48±0.14 | 1.80±0.16 | <0.001 |
| TyG | 8.45±0.69 | 8.34±0.68 | 8.58±0.67 | <0.001 |
| Income to Poverty Ratio | 2.61±1.56 | 2.64±1.58 | 2.58±1.54 | 0.291 |

Values are Mean ±SD (for datas with normal distribution) and n (%).
BMI=Body Mass Index, W/H=Waist/Hipline, SBP=Systolic Blood Pressure, DBP=Diastolic Blood Pressure, MAP=Mean Atrial Pressure, TG=Triglyceride, CHOL=Total Cholesterol, LDL=Low Density Lipoprotein, HDL=High Density Lipoprotein, GHB=Glycohemoglobin, GLU=Glucose, LAP=Lipid Accumulation Product, VAI=Visceral Adiposity Index, TyG=Triglyceride-Glucose Index.

### Principal Component Analysis

PCA was employed for data dimensionality reduction. Figure 2A illustrates the variance and cumulative variance curve of every PCs among all participants. The first 6 PCs accounted for 85% of the variance. The contributions of each independent variable to PC1 to PC6 are shown in Figure 2B. |Loading value| of variables exceeding 0.6 were retained as the indicators for hierarchical clustering, including obesity (Height, Weight, BMI, Waist, Hipline, W/H), blood pressure, blood lipids (CHOL, TG. LDL, HDL), and IR (GHB, GLU, VAI, LAP, TyG) indicators.

**Figure 2.**
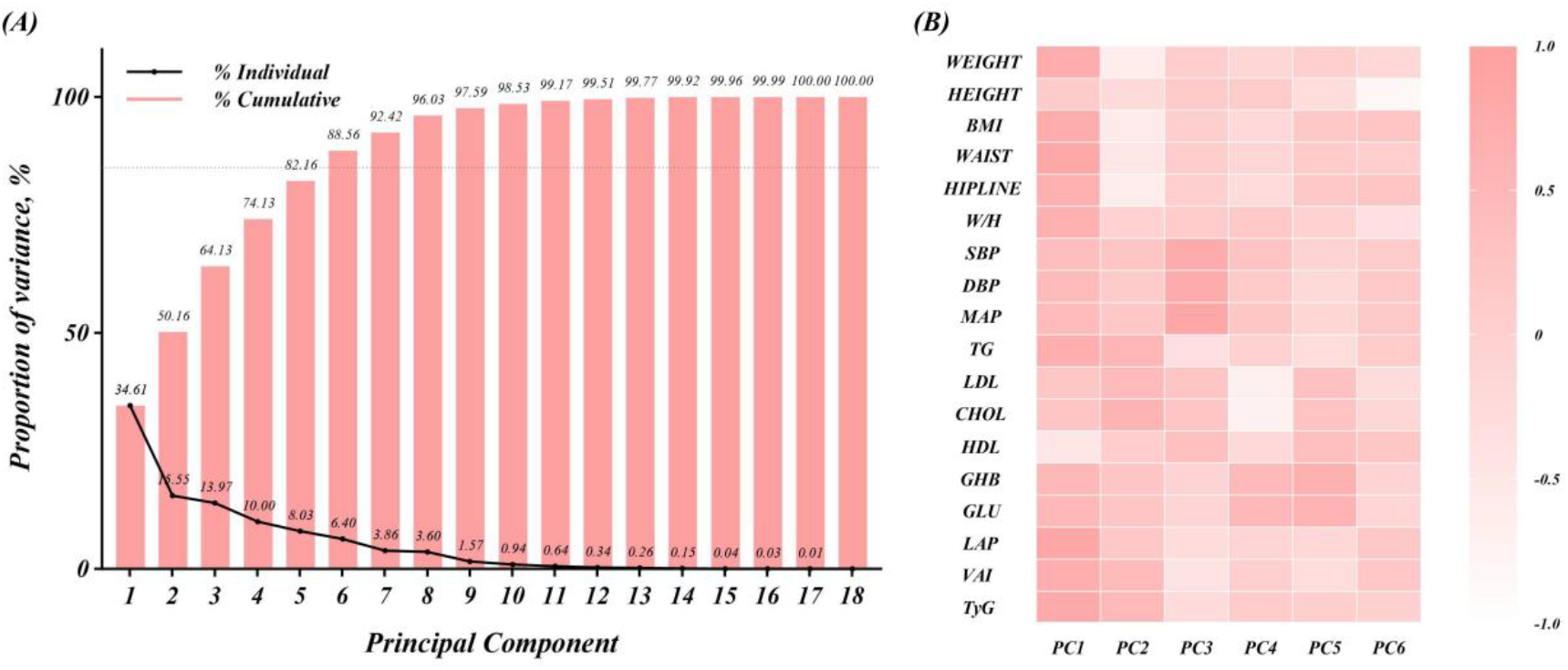
PCA of all participants. **(A)** Variance explained of individual PC and cumulative variance. PCs explain for a cumulative variance of 85% were used for further analysis. **(B)** Loading plot shows the loading values of each variable on a single PC. Variables with a |loading value|≥ 0.6 were included in the cluster analysis.

Similarly, Figures 3A and Figure 3B show the variance and cumulative variance curve of every PCs and the contributions of independent variables to PCs among OSA participants. The indicators for hierarchical clustering among OSA participants included obesity (Height, Weight, BMI, Waist, Hipline, W/H), blood pressure, blood lipids (CHOL, TG. LDL, HDL), and IR (GHB, GLU, VAI, LAP, TyG) indicators.

**Figure 3.**
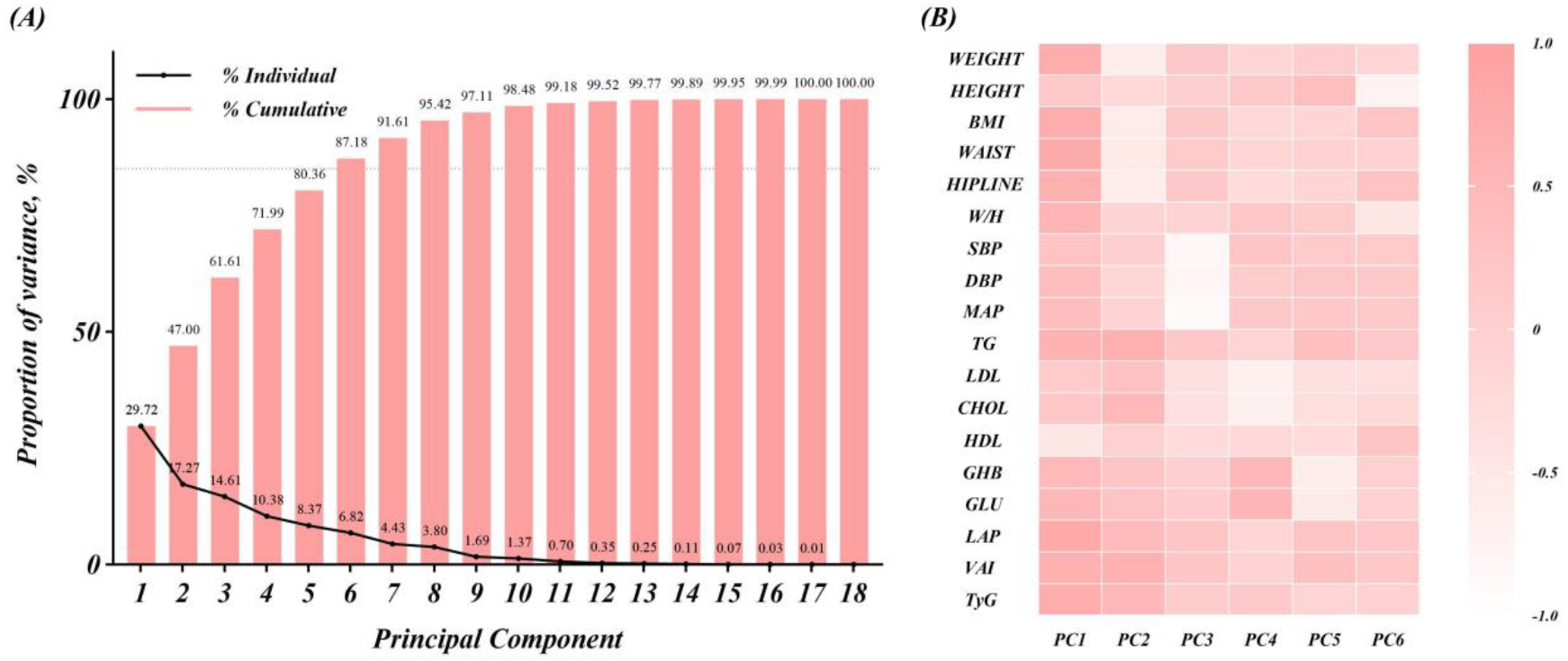
PCA of OSA participants. **(A)** Variance explained of individual PC and cumulative variance. **(B)** Loading plot shows the loading values of individual variable on a single PC. Others are presented in Figure 2.

### Hierarchical Clustering

The hierarchical clustering process is shown in Supplementary Figure 2. To better identify the differences among clusters, a relatively large number of clusters were chosen[11]. The overall participants were divided into 6 clusters, while the OSA participants were divided into 8 clusters.

### Characteristics of Clusters

Among all the participants (Figure 4, SupplementaryTable 1), Cluster 1 (Figure 4A) exhibited the lowest OSA prevalence (374/1136, 32.92%), whose level of obesity, blood pressure, IR, and blood lipid levels are superior to population averages. Cluster 2 (Figure 4B) demonstrated elevated OSA incidence (166/291, 57.04%), associated with marked elevations in IR markers, as well as an increase in blood pressure and blood lipid levels, while the degree of obesity is not significantly increased. In Clusters 3 (226/395, 57.21%) (Figure 4C) and Cluster 4 (547/883, 61.94%) (Figure 4D), the OSA levels of participants are also higher than the average level. Cluster 3 is mainly characterized by a significant increase in the degree of obesity, while the levels of other metabolic indicators are almost consistent with the average level. In Cluster 4, participants mainly show a mild increase in the degree of obesity and blood pressure levels. The incidence of OSA in Clusters 5 (155/325, 47.69%) (Figure 4E) and Cluster 6 (221/435, 50.80%) (Figure 4F) is almost consistent with the overall OSA incidence rate, but their degree of obesity is lower than the overall level. Cluster 5 is mainly characterized by an increase in CHOL and HDL levels (Figure 4E), while Cluster 6 is mainly characterized by an increase in blood pressure levels (Figure 4F).

**Figure 4.**
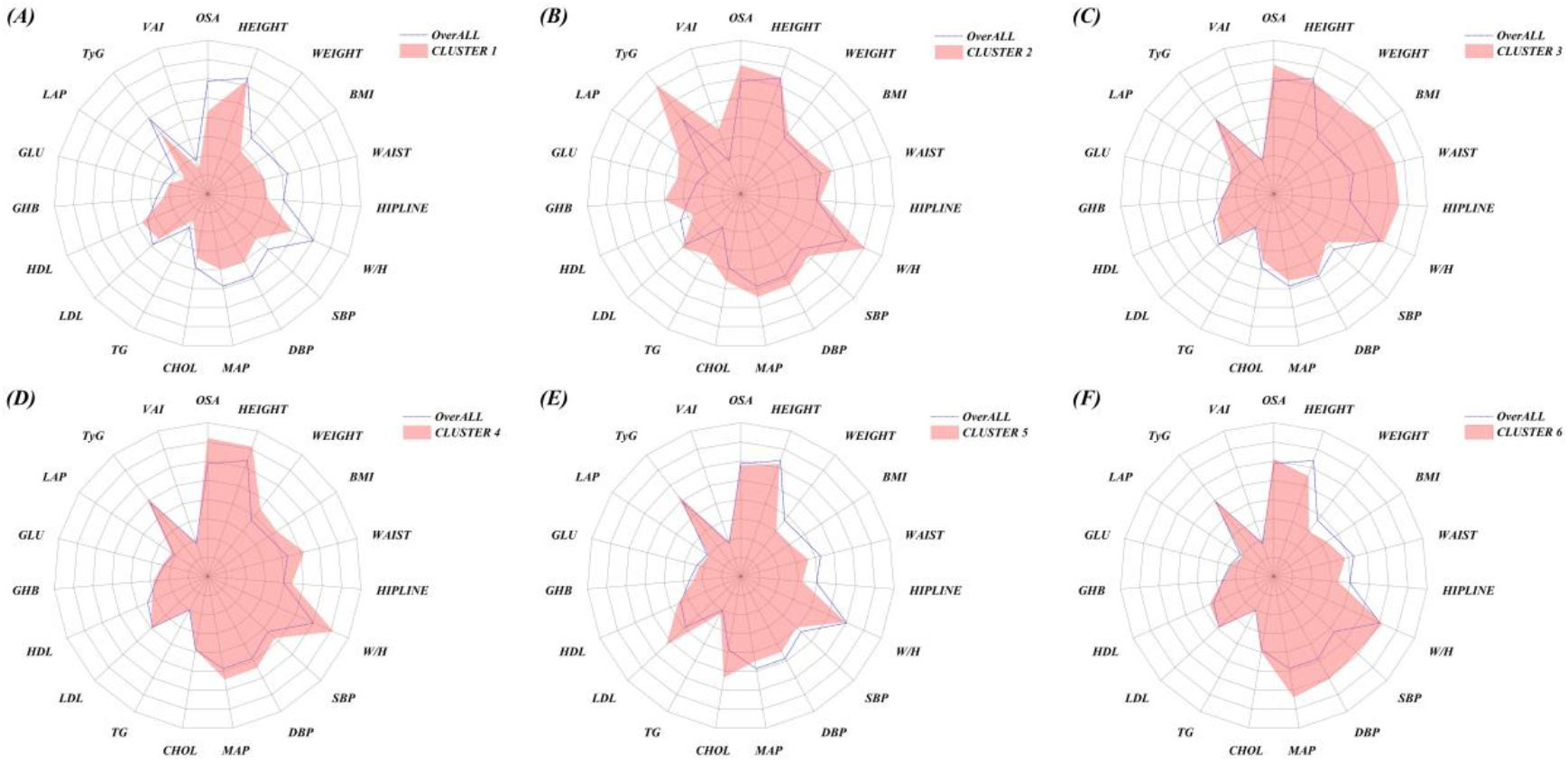
Distribution of metabolic index of all participants. **(A) - (F)**. Radar chart displays the distribution of individual metabolic index in every cluster. The dashed line represents the mean of metabolic index of all participants, the red pattern represents the mean of metabolic index of every cluster. The mean values of metabolic index were subjected to Min-Max Normalization before the radar chart was plotted.

Among the OSA participants, different OSA sub-phenotypes were also identified (Figure 5, Supplementary Table 2). Cluster A (Figure 5A) participants were mainly characterized by increased blood pressure levels and decreased obesity levels. Cluster B (Figure 5B) was characterized by elevated HDL and reduced levels of all other metabolic indicators. Cluster C (Figure 5C) was characterized by decreased obesity and blood pressure levels, and increased CHOL and LDL levels. Cluster D (Figure 5D) was mainly characterized by decreased blood pressure levels, with other metabolic indicators being consistent with the overall OSA participants. Cluster E (Figure 5E) was characterized by a significant increase in obesity levels, with no significant changes in other metabolic indicators. Cluster F (Figure 5F) also showed a significant increase in obesity levels, accompanied by elevated blood pressure, and its main difference from Cluster E (Figure 5F) was in the levels of LDL and GHB. Clusters G (Figure 5G) and H (Figure 5H) had obesity levels consistent with the average level, but with significantly elevated insulin resistance indicators.

**Figure 5.**
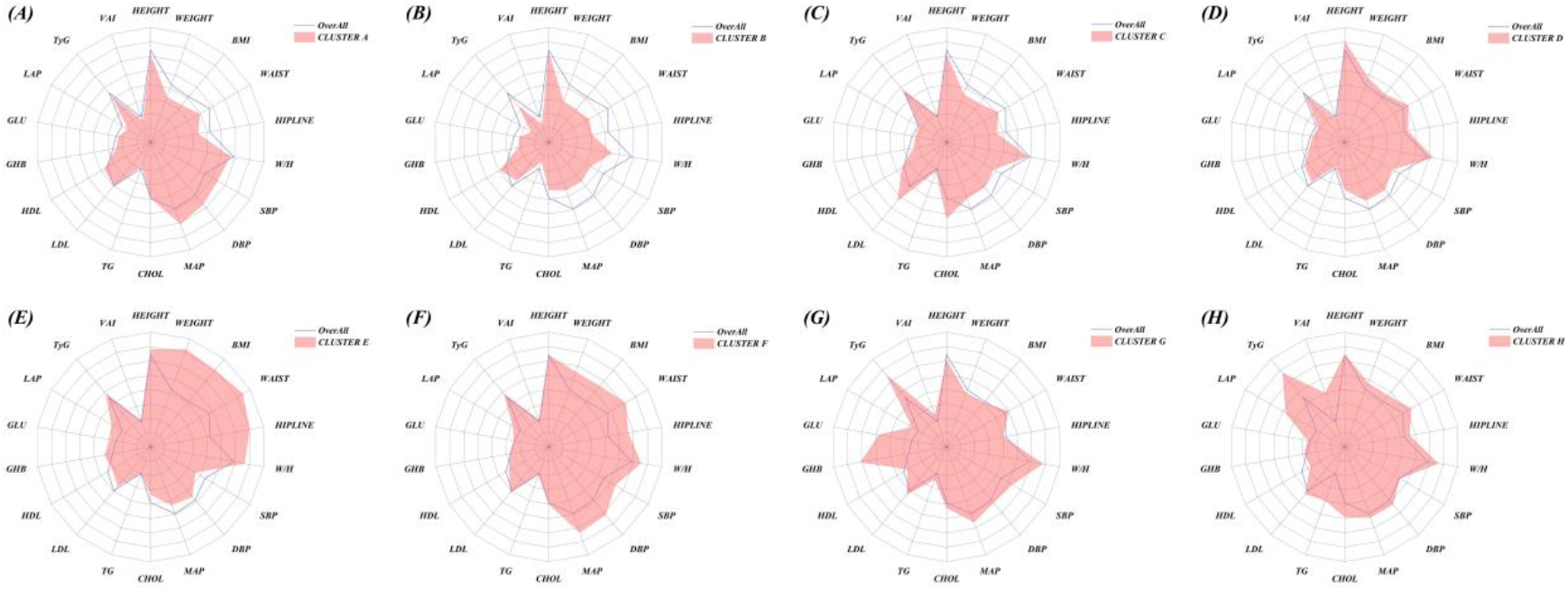
Distribution of metabolic index of OSA participants. **(A) - (H)**. Radar chart displays the distribution of individual metabolic index in every cluster. The dashed line represents the mean of metabolic index of OSA participants. Others are presented in Figure 4.

## Discussion

The present study leveraged NHANES data to dissect the heterogeneous metabolic profiles associated with OSA, revealing distinct subphenotypes driven by varying contributions of obesity, IR, blood pressure, and lipid abnormalities. These findings underscore the complexity of the OSA-MetS relationship and highlight the potential for targeted interventions based on metabolic drivers.

### Duality of Obesity Indicators

In the entire cohort, the obesity indicators in Cluster 3, where the prevalence of OSA is above average, are significantly higher than the average level. These results are consistent with prior evidence highlighting obesity as a central risk factor for OSA[12, 13]. However, the W/H in this group is similar to the average level, indicating that this group does not represent a population with obvious central obesity, more likely a subtype of isolated obesity participants. Similarly, the characteristics of Cluster E in OSA participations are also marked by isolated obesity, with W/H and other indicators not exceeding the average level. We think that the primary mechanism by which obesity contributes to OSA in these clusters is likely due to airway narrowing and reduced lung volumes caused by fat accumulation[14]. Additionally, endocrine abnormalities associated with obesity can lead to autonomic nervous dysfunction[15], thereby exacerbating nocturnal airway obstruction and reduced lung volumes[16]. This is because an almost completely different phenomenon was observed in Cluster 2 in all participants, where the prevalence of OSA is also above average. In Cluster 2, the obesity indicators are similar to the average level, while indicators such as IR, dyslipidemia, and hypertension are all significantly elevated, and the W/H is also above average. It is evident that Cluster 2 may represent a completely different category from Cluster 3, namely participants with metabolic abnormalities but without apparent obesity. Similar participants were also identified in Clusters G and H among OSA participants. Researchers have found that IR and other metabolic abnormalities occur in animal models of OSA or CIH and healthy subjects[17-19]. It has been shown that systemic inflammatory responses caused by central obesity are more pronounced[20]. This suggests the existence and rationality of non-uniform metabolic abnormality subtypes of obesity/non-obesity among OSA patients. Therefore, we propose the hypothesis that the role of obesity in the occurrence and progression of OSA is determined by the type of obesity and the presence of associated metabolic abnormalities. Moreover, this also provides evidence for the existence of metabolically healthy obesity and metabolically abnormal normal weight.

### IR in Obese and Non-Obese OSA

In Cluster 2 of all participants, where the prevalence of OSA is also above average, participants exhibited a general increase in metabolic indicators other than obesity, with a particularly significant elevation in IR indicators. Similarly, Clusters G and H were primarily characterized by a marked increase in IR markers. This suggests that IR may play a significant role in the development and progression of non-obese OSA. The development of IR and pancreatic beta cell dysfunction has been linked to CIH in OSA[21]. CIH exacerbated fasting hyperglycemia, glucose intolerance, and IR in both mice with diet-induced obesity and mice with genetic obesity[22-24]. Apart from obese mice, CIH-induced IR could also be observed in lean vivo and healthy volunteers[25, 26]. This confirms that OSA can interact with IR independently of obesity and also identifies a susceptible population of OSA characterized by isolated IR/diabetes.

Besides, it is worth noting that participants in Cluster 2 of all participants not only had elevated IR indicators but also had higher waist circumference and W/H than the average level. As we all know, LAP/VAI not only reflect the degree of IR but are also directly related to the accumulation of visceral fat, which may exacerbate OSA through pro-inflammatory cytokines (e.g., TNF-α, IL-6)[27, 28] that reduce upper airway muscle tone[29]. This creates a vicious cycle where OSA worsens IR, which in turn aggravates airway collapsibility. In the analysis of participants with OSA, Cluster G and Cluster H correspond to the characteristics of this special group. This confirms that OSA can interact with IR by isolated visceral fat accumulation ranther than obvious obesity and also identifies a susceptible population of OSA characterized by isolated IR/diabetes.

### Heterogeneity of Hypertension in OSA

Clustering analysis also revealed an interesting phenomenon regarding another component of MetS—hypertension. This could be observed in Cluster 6 of all participants. This is inconsistent with our usual understanding, as hypertension typically coexists with obesity in OSA patients[30, 31], and even the blood pressure levels of the obese participants in Cluster 3 were lower than the average level. In OSA participants, Cluster A also showed the same pattern. This may explain the inconsistent efficacy of CPAP treatment for OSA-related hypertension[32]. OSA subgroups characterized by isolated hypertension may be more responsive to CPAP therapy, as their blood pressure levels are less influenced by metabolic abnormalities. This hypothesis is supported by the fact that patients with hypertension combined with metabolic abnormalities are less sensitive to antihypertensive treatments compared to those with isolated essential hypertension[32, 33]. Animal experiments have also demonstrated that isolated CIH can increase carotid body sensitivity, leading to sustained adrenergic drive and subsequently causing hypertension[34, 35]. Moreover, CIH can reduce the bioavailability of nitric oxide, leading to endothelial dysfunction and consequently hypertension[36, 37]. For these OSA models with hypertension without metabolic abnormalities, high-concentration oxygen therapy can alleviate elevated blood pressure. Therefore, we believe that it is necessary to differentiate between metabolic-normal hypertension and metabolic-abnormal hypertension in OSA patients, as the mechanisms underlying their hypertension may be entirely different, and their sensitivity to various antihypertensive treatments may also vary. This requires confirmation through larger cohort studies.

### Study Limitations

Firstly, the clinical diagnosis of OSA is currently based on sleep monitoring tests. In this study, we adopted a relatively lenient diagnostic criterion for OSA as used in previous studies[7], which may have overestimated the prevalence of OSA in the population. Secondly, although the NHANES database provides detailed metabolic indicators, we were still unable to identify a direct measure representing IR. Thirdly, we attempted to merge data from multiple NHANES survey cycles; however, after data cleaning, the sample size did not increase significantly. This may be due to inconsistencies in certain variables across different survey cycles. Therefore, we conducted our analysis using only the 201–2020 (pre-pandemic) data. Lastly, due to workload constraints, we did not conduct an in-depth analysis of the characteristics of OSA participants in each metabolic subtype, which will be addressed in future studies.

## Conclusion

In conclusion, our analysis of NHANES data reveals substantial metabolic heterogeneity among individuals with OSA, characterized by distinct subphenotypes with differing profiles of obesity, IR, hypertension, and dyslipidemia. These findings suggest that OSA may involve multiple metabolic interaction pathways, underscoring the need for personalized treatment strategies tailored to specific metabolic profiles.

## Supplementary Material

Supplementary material is available at Circulation online.

## Acknowledgements

We thank than the following persons for help during the study. FKX and ZJX designed the study. FKX, ZXF performed the experiments. FKX, SJK, XBW, DHB analyzed the data. FKX, ZJX, YJ, GLand ZL prepared the manuscript. All authors have seen and approved the final version of this manuscript.

## Data availability

The data underlying this article will be shared on reasonable request to the corresponding author.

